# Alterations in Brain Morphology by MRI in Adults with Neurofibromatosis 1

**DOI:** 10.1101/2019.12.19.19015255

**Authors:** Su Wang, Victor-Felix Mautner, Ralph Buchert, Stephane Flibotte, Per Suppa, Jan M. Friedman, Manraj K. S. Heran

## Abstract

**Objective:** To characterize alterations in brain morphology by MRI in adults with neurofibromatosis 1 (NF1).

**Methods:** Planar (2D) MRI measurements of 29 intracranial structures were compared in 389 adults with NF1 and 112 age- and sex-matched unaffected control subjects. The 2D measurements were correlated to volumetric (3D) brain measurements for 99 of the adults with NF1.

A subset of adults with NF1 (n = 70) was also assessed for clinical severity of NF1 features and neurological problems and received psychometric testing for attention deficits and IQ. Correlation analyses were performed between principal components of the intracranial measurements and clinical and psychometric features of these patients.

**Results:** Four of nine corpus callosum measurements were significantly greater in adults with NF1 than in sex- and age-matched controls. All seven brainstem measurements were significantly greater in adults with NF1 than in controls. Increased corpus callosum and brainstem 2D morphology were correlated with increased total white matter volume among the NF1 patients. No robust correlations were observed between the 2D size of these structures and clinical or neuropsychometric assessments.

**Interpretation:** Our findings are consistent with the hypothesis that dysregulation of brain myelin production is an important manifestation of NF1 in adults.

## INTRODUCTION

Neurofibromatosis 1 (NF1), an autosomal dominant disease caused by mutations of the *NF1* gene, affects approximately 1 in 3000 live births^1-3^. NF1 causes a wide range of clinical features: neurofibromas, café-au-lait macules, macrocephaly, learning disabilities, and attention deficits^4-15^. Associated gliomas, malignant peripheral nerve sheath tumours, skeletal dysplasias, and cardiovascular disease may cause serious disability or death in affected children or young adults. The *NF1* gene encodes neurofibromin, which is a negative regulator of the RAS cellular proliferation pathway^16^. Loss of NF1 function results in increased Schwann cell proliferation, a primary feature of NF1 pathology^16^.

Many studies have used magnetic resonance imaging (MRI) to study brain morphology in children and adults with NF1. NF1 neuroimaging studies have been especially focused on unidentified bright objects, optic nerve/chiasmatic expansions, and tumours of the peripheral nerves or central nervous system^17, 18^. Most people with NF1 do not develop brain tumours but may exhibit other, more subtle alterations of brain morphology on MRI examination, including increases in total white matter volume, total brain volume, corpus callosum (CC) area, CC length, and optic nerve tortuosity (ONT)^6, 7, 9, 10, 19-28^. The clinical implications, if any, of these findings are unclear, given the small number and young age of individuals with NF1 in most reported studies^10^.

Previous research has attempted to correlate brain morphological differences with cognitive or behavioural abnormalities in children with NF1, but inconsistent results have been observed^10, 24, 29, 30^. To date, there have been no large-scale MRI studies of brain morphological differences and their possible relationship to cognitive or behavioural abnormalities in adults with NF1.

We characterized 2D brain morphological differences in adults with NF1 within three regions of interest: the CC, brainstem, and optic nerves and ocular globes. We correlated the 2D measurements for the CC and brainstem with 3D volumetric measurements in NF1 to assess the role of different brain composition in the structural changes. Finally, we examined the associations of these 2D brain morphological changes with neuropsychometric findings among adults with NF1.

## METHODS

### Participants

Between 2003 and 2015, all patients seen in the NF Outpatient Department of the University Hospital Hamburg-Eppendorf in Hamburg, Germany, were offered brain MRI to monitor their intracranial tumour burden. Head MRI was obtained on 434 adults with NF1; each patient was imaged an average of three times. Participants were excluded from the current study if they had optic gliomas or brain tumors. Sex and age were recorded at the time of each examination.

Unaffected control individuals, who had undergone head MRI for other reasons at Vancouver General Hospital, Vancouver, Canada, were age- (within 24 months) and sex-matched to 25% of randomly selected adults with NF1. Affected and unaffected participants were excluded if no image was available that permitted clearly defined morphological measurements. In total, 389 adults with NF1 and 112 unaffected controls were included in the morphological analysis. In cases where multiple MRI scans were obtained for one individual, the latest MRI scan was used for brain morphological measurements.

A subset of adults with NF1 (n = 70) received clinical evaluations for NF1 severity, neurological severity, attention deficit hyperactivity disorder (ADHD) severity, and neuropsychometric evaluations for attention deficiencies and intelligence quotient (IQ).

### Standard Protocol Approvals, Registrations, and Patient Consent

The ethical committees of the Medical Chamber in Hamburg and the Research Ethics Board of the University of British Columbia approved the study. Written consent was obtained from all study participants before the study began. All data were de-identified before analysis.

### Morphological Measurements

Head MRI exams were conducted in a 1.5 or 3.0 Tesla scanner. Multiple coronal, axial, and/or sagittal T1, T2, and FLAIR images were obtained. OsiriX Lite^31^ software was used to obtain measurements in NF1 patients, and IMPAX PACS (Agfa, Ridgefield Park, NJ) software was used for this purpose in unaffected controls. Image slices were ≤ 5.5 mm thick for adults with NF1 and ≤ 6 mm thick for unaffected populations. In total, 29 measurements were obtained for each MRI set for each participant.

Eight measurements were obtained for the CC: area, length, height, genu width, anterior body width, mid-body width, posterior body width, and splenium width (Figure 1A). CC height was measured as maximum height of CC perpendicular to a line drawn at base of the CC.

**Figure 1:**
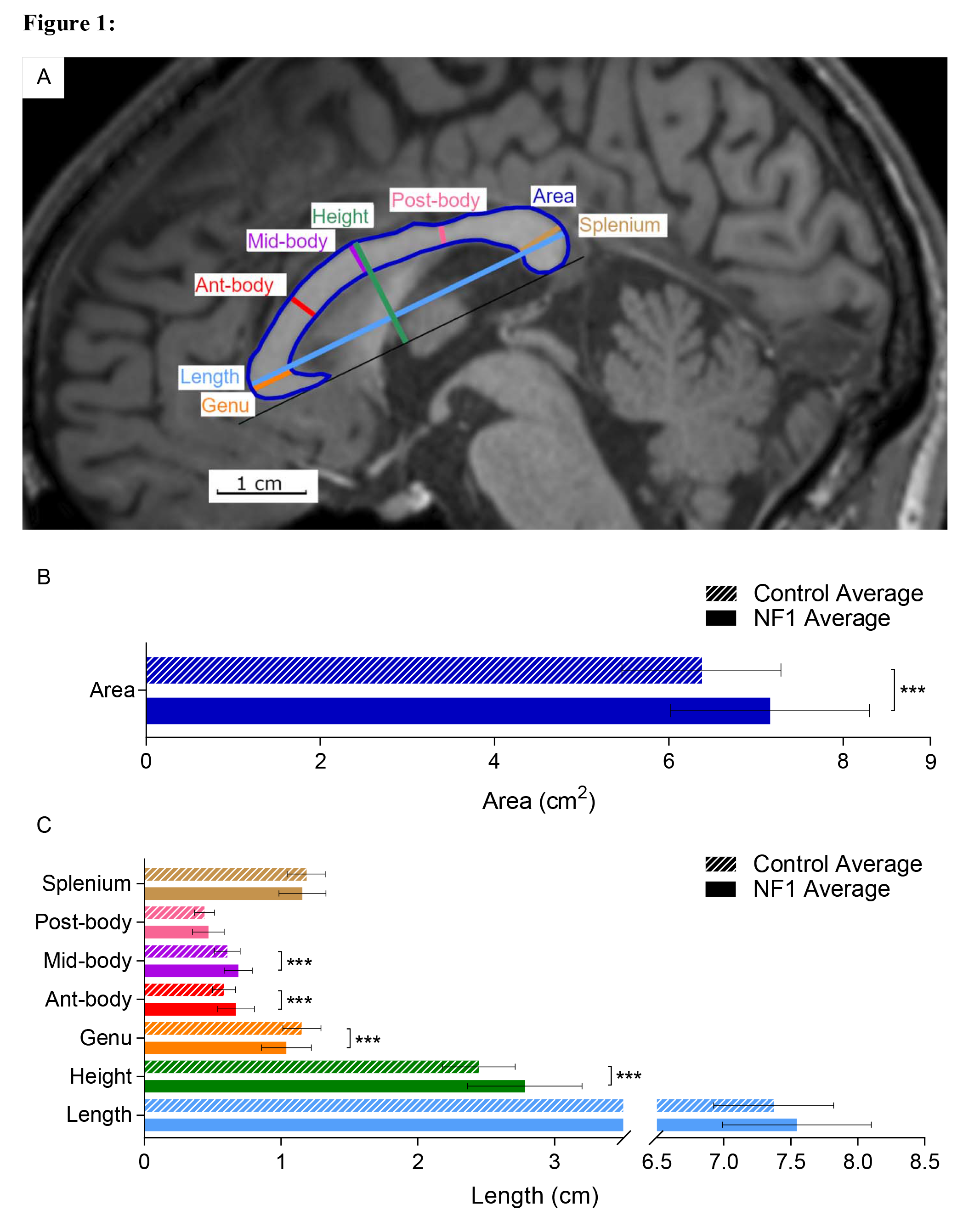
Corpus callosum measurements and averages for adults with NF1 and unaffected control participants. (A) Representative MRI image showing T1-weighted midline sagittal view of a male in his 30’s with NF1. (B) Average CC midsagittal area. (C) Average CC measurements. Coloured lines and area outlined in (A) show where structures indicated by the same colour in (B) or (C) are measured. Error bars indicating one standard deviation and asterisks indicating FDR-adjusted statistical significance are shown in (B) and (C). p < 0.001 =***. Abbreviations used in figure: Genu = genu width, Ant-body = anterior body width, Mid-body = mid-body width, Post-body = posterior body width, and Splenium = splenium width.

Seven measurements were obtained for each of the ocular globes: anterior to posterior (AP) length, diameter, anterior to diameter (AD) length, anterior to interzygomatic line (AIZ) length, posterior to interzygomatic line (PIZ) length, optic nerve displacement (OND), and optic nerve path (ONP) (Figure 2A). AP and diameter measurements were obtained at maximum length/width. AD was measured as maximum length perpendicular to diameter. AIZ and PIZ was measured as maximum length perpendicular to interzygomatic line.

**Figure 2:**
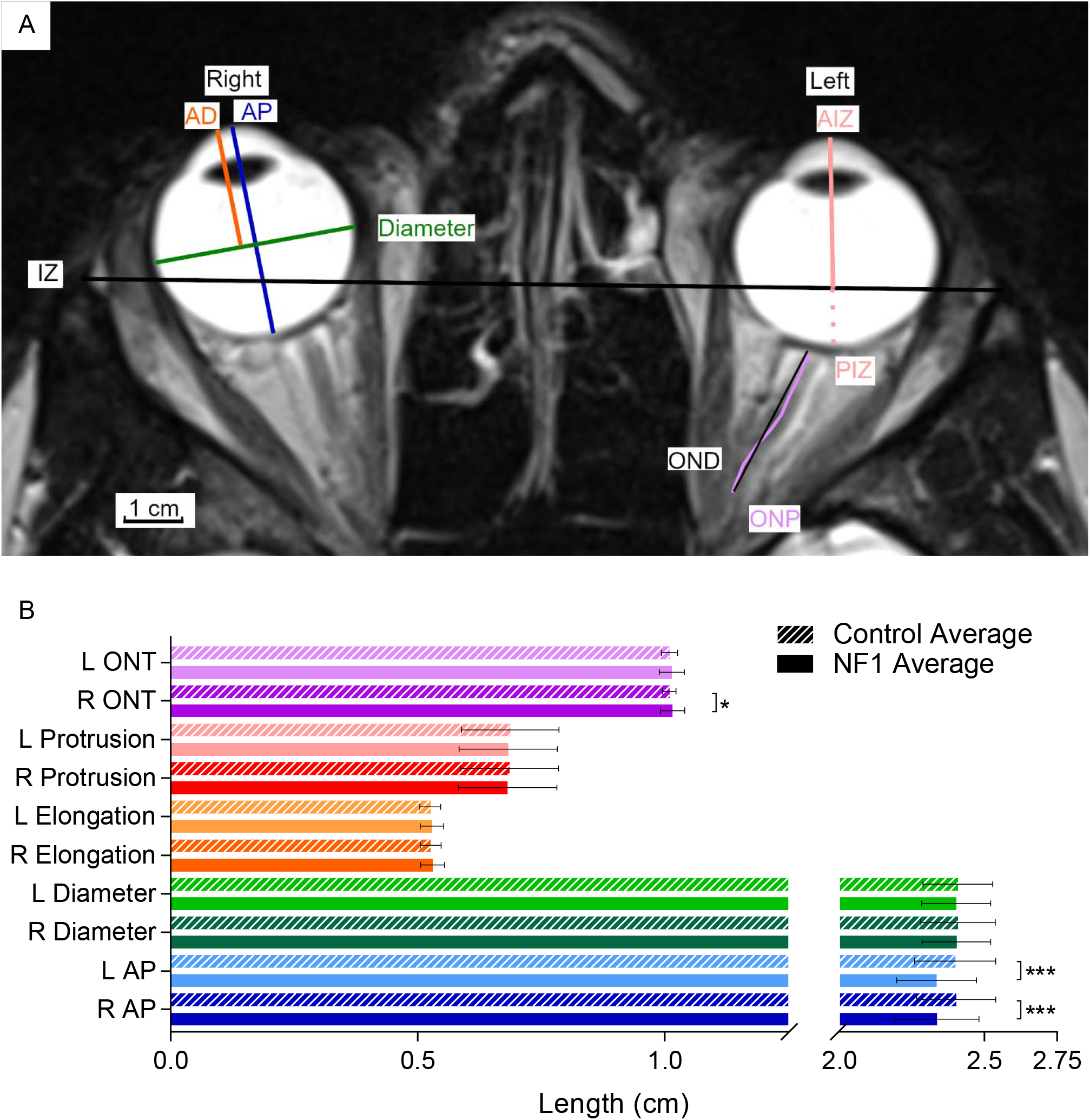
Ocular globe measurements and averages for adults with NF1 and unaffected control participants. (A) Representative MRI image showing T2-weighted axial view of a male in his 20’s with NF1. (B) Select average ocular globe measurements. Unique measurements are shown on one side but were obtained on both left and right side. Coloured lines in (A) show where structures indicated by the same colour in (B) are measured. Error bars indicating one standard deviation and asterisks indicating FDR-adjusted statistical significance are shown in (B). p < 0.5 = *; p < 0.001 = ***. Abbreviations used in the figure: L = left, R = right, AP = anterior to posterior length, AD = anterior to diameter length, AIZ = anterior to interzygomatic line length, IZ = interzygomatic line, PIZ = posterior to interzygomatic line length, OND = optic nerve displacement, and ONP = optic nerve path.

Seven measurements were obtained for the brainstem: midbrain width, midbrain AP length, pons AP length, left and right middle cerebellar peduncle (MCP) lengths, medulla oblongata width, and medulla oblongata AP length (Figure 3A). Midbrain width, pons AP, left and right MCP, and medulla oblongata width were measured as maximum length/width. Midbrain AP and medulla oblongata AP length were measured as maximum length perpendicular to midbrain width and medulla oblongata width, respectively.

**Figure 3:**
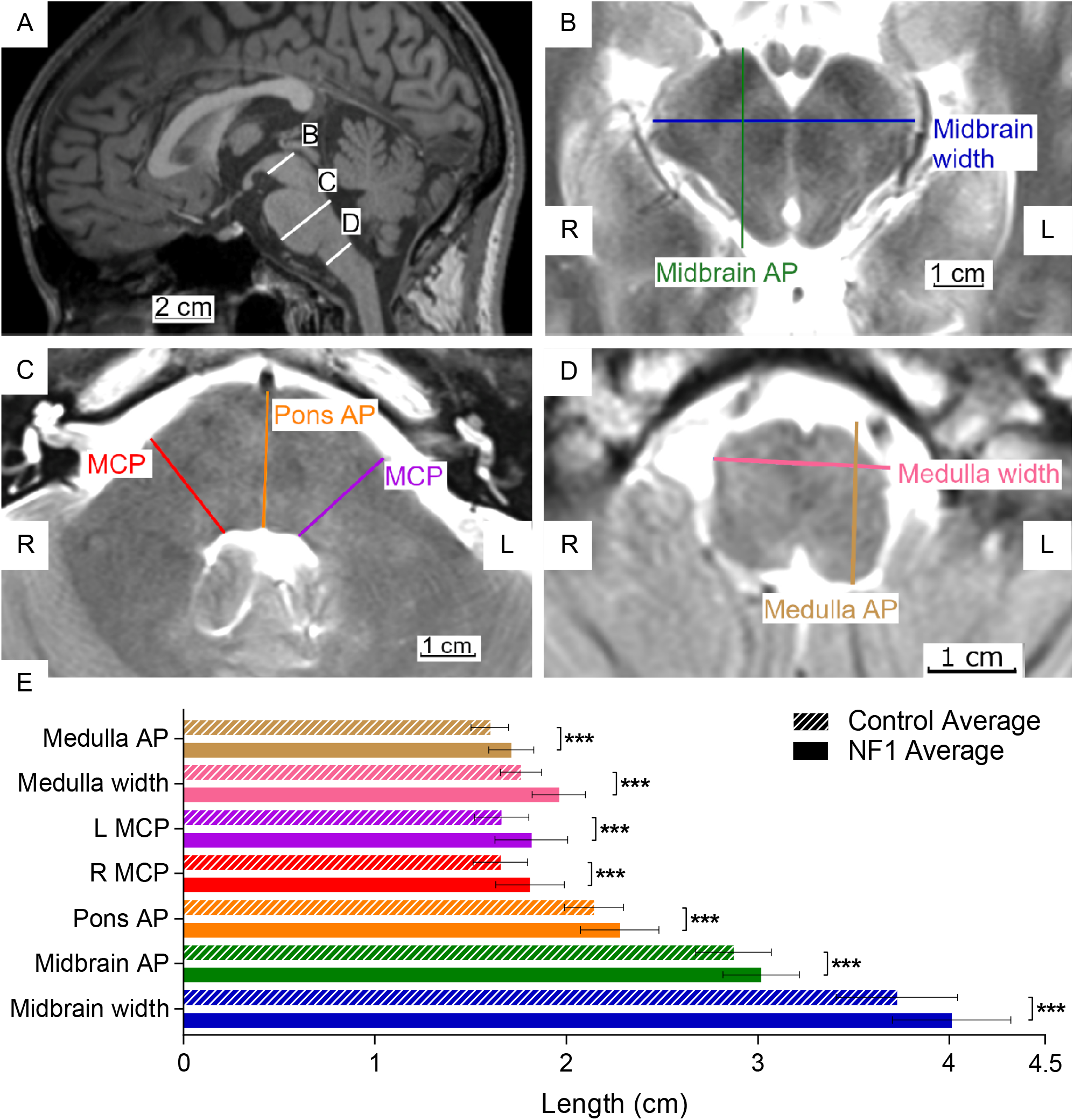
Location of brainstem measurements shown on representative sagittal (A), axial (B), (C), and (D) images, and average measurements (E) for adults with NF1 and unaffected control participants. (A-D) Representative MRI images showing T1-weighted midline sagittal and T2-weighted axial views of a male 20’s with NF1. (E) Average brainstem measurements. The labeled lines in (A) show position of the midbrain (B), the pons and the middle cerebellar peduncle (C), and the medulla oblongata (D). Coloured lines in (B-D) show where structures indicated by the same colour in (E) are measured. Error bars indicating one standard deviation and asterisks indicating FDR-adjusted statistical significance are shown in (E). p < 0.001 = ***. Abbreviations used in the figure: L = left, R = right, AP = anterior to posterior length, and MCP = middle cerebellar peduncle.

CC bulbosity was calculated by dividing the splenium width by the posterior body width. Ocular globe elongation was calculated by dividing AD length by AP length. Globe protrusion was calculated by dividing AIZ by the sum of AIZ and PIZ. Optic nerve tortuosity (ONT) was calculated by dividing ONP by OND.

Brain volumes were measured according to methods described in our companion paper^28^.

### Clinical and Neuropsychometric Assessments

#### Clinical NF1 Severity

Clinical severity for adults with NF1 (n = 70) was categorised into four grades by Dr. Victor Mautner using a modified Riccardi scale^32, 33^. Intellectual and psychological functions were excluded from the NF1 clinical severity rating to avoid confounding between learning disability and medical severity.

#### Clinical Neurological Severity

Clinical neurological severity for adults with NF1 (n = 70) was categorised by Dr. Victor Mautner into one of four grades. Grade 1 encompasses no deficits. Grade 2 includes discrete neurological deficits only: muscle hypotonia, sensation deficits, balance problems, or speech problems. Grade 3 includes neurological dysfunction: paresis, ataxia, significant ocular motor deficits, or substantial pain. Grade 4 encompasses neurological deficits that seriously compromise health: intractable pain, paralysis, or drug-resistant seizures.

#### Clinical Attention deficit hyperactivity disorder (ADHD) Severity

Clinical ADHD severity for adults with NF1 (n = 50) was categorized into three grades using the established criteria in DSM-IV^34^. Grade 1 encompasses no clinical diagnosis. Grade 2 is subclinical attention deficit with no ADHD diagnosis. Grade 3 includes those with a diagnosis of either Attention Deficit Disorder (ADD) or ADHD.

#### Intelligence quotient (IQ)

Full-scale, verbal, and performance IQ were obtained for 61 adults with NF1 using the Wechsler Adult Intelligence Scale-Revised (WAIS-R)^35^. WAIS-R test scores were standardized according to age and sex.

#### Attention Deficit Measurement

Attention deficit measurements for adults with NF1 (n = 68) were obtained using the Visual Test of Variables of Attention, version 7.0.3 or 8.0^36^. Measurements were obtained for variability of response time (consistency), response time, commission error (impulsivity), errors of omission (inattention), post-commission response times, and multiple and anticipatory responses. An Attention Comparison Score was calculated for each patient. All test scores were standardized according to age and sex.

### Statistical Analysis

All data were analysed using R Studio 3.4.1^37^. A False Discovery Rate (FDR) adjusted significance level of p < 0.05 was used throughout to account for multiple comparisons^38^.

#### Demographic Analysis

Mean age was compared between adults with NF1 and unaffected controls using the Student-T test after demonstrating satisfactory dataset normality and variance using the Shapiro-Wilk test and Fisher’s F test, respectively. Sex ratios were compared between adults with NF1 and unaffected controls using the χ^2^ test.

#### Brain Morphological Analysis

Means of the 26 different brain morphological measurements were compared between adults with NF1 and unaffected controls. Student-T tests were used for normally distributed data. Non-normal distributions were compared using Mann-Whitney U tests. CC bulbosity means were compared between males and females for both adults with NF1 and unaffected adults using two-way ANOVA test. Dataset normality and variance were determined using the Shapiro-Wilk test and Fisher’s F test respectively. Significance (p-value) was adjusted using FDR to account for 26 comparisons.

#### Brain Morphological – Volumetric Correlation Analysis

Brain morphological – volumetric correlation analysis was only conducted for adults with complete 2D CC or brainstem measurements and 3D volumetric measurements. Principal component analysis (PCA) was calculated for CC and brainstem separately. PCs were included based on a combination of eigenvalue (greater than 1) and minimum combined amount of variance it accounts for (70%). Pearson correlation was conducted, and p values were adjusted using FDR to account for multiple comparisons.

#### Brain Morphology – Neuropsychometric Correlation Analysis

Brain morphology measurements for the CC, ocular globes, and brainstem were independently grouped using PCA to maximize the number of subjects included in the PCA. PCs for the correlation analysis were selected based on same threshold as above described. Clinical NF1 severity, clinical neurological severity, and clinical ADHD diagnosis data were analysed as ordinal data. Attention Comparison Score was analysed as a continuous measurement of attention deficit. Total IQ scores from WAIS-R were analysed as a continuous measurement. PCs from the three brain structures were independently compared to the neuropsychometric measurements using Pearson correlation for continuous data and Spearman’s Rank-Order correlation for ordinal data. No significance testing was conducted on the correlation matrix due to insufficient sample size.

### Data Availability

Anonymous data are available for appropriate research purposes through V.F. Mautner, MD.

## RESULTS

### Demographics

This study included 494 participants: 389 adults with NF1 and 112 age- (within 24 months) and sex-matched unaffected controls. There were no statistically significant differences in age or sex ratio distribution between adults with NF1 and unaffected controls overall or in the subgroups in whom the three brain structures (CC, ocular globes, and brainstem) were measured by MRI (Table 1).

**Table 1:** Participant demographics for adults with NF1 and matched unaffected controls.

|  | ADULTS WITH NF1 | UNAFFECTED CONTROLS |
| --- | --- | --- |
| ALL SUBJECTS |  |  |
| N | 389 | 112 |
| Mean Age (SD) | 37.4 (13.5) | 39.4 (13.1) |
| Age Range | 18.1 - 72.9 | 19.4 - 73.3 |
| Female: Male<br>(Female Percentage) | 218:171 (56.0%) | 63:49 (56.2%) |
| SUBJECTS WITH MRI MEASUREMENTS OF CORPUS CALLOSUM |  |  |
| N | 226 | 57 |
| Mean Age (SD) | 37.1 (13.6) | 38.0 (13.2) |
| Age Range | 18.2 - 72.9 | 19.4 - 64.7 |
| Female: Male<br>(Female Percentage) | 126:100 (55.7%) | 31:26 (54.4%) |
| SUBJECTS WITH MRI MEASUREMENTS OF OCULAR GLOBES |  |  |
| N | 386 | 96 |
| Mean Age (SD) | 37.4 (13.5) | 39.6 (13.7) |
| Age Range | 18.1 - 72.9 | 19.4 - 73.3 |
| Female: Male<br>(Female Percentage) | 214:172 (55.4%) | 56:40 (58.3%) |
| SUBJECTS WITH MRI MEASUREMENTS OF BRAIN STEM |  |  |
| N | 375 | 96 |
| Mean Age (SD) | 37.2 (13.3) | 39.5 (13.1) |
| Age Range | 18.1 - 72.9 | 19.4 - 73.3 |
| Female: Male<br>(Female Percentage) | 209:166 (55.7%) | 56:40 (58.3%) |

The demographics are analysed in total and in measurement groups. No comparisons between adults with NF1 and unaffected controls were significantly different.

### Brain Morphology Comparison Between NF1 Patients and Controls

Four of eight corpus callosum measurements (midsagittal length, height, anterior body width, mid body width) were significantly greater and one (genu width) was significantly shorter in adults with NF1 than in unaffected control participants (Figure 1B, C). CC length, posterior body width, splenium width, and CC bulbosity did not differ between the two groups. The statistical significance was determined after adjustment for multiple comparisons.

Of the five measurements made for each ocular globe, only the AP lengths were significantly reduced in both eyes in adults with NF1 compared to unaffected controls (Figure 2B). Optic nerve tortuosity was greater among adults with NF1 than among unaffected individuals. This difference was small and reached statistical significance only on the left side.

The brainstems of individuals with NF1 were significantly larger than expected in all seven sites measured (midbrain width, midbrain AP length, pons AP length, left and right MCP lengths, medulla oblongata width, and medulla oblongata AP length) (Figure 3E).

### Brain Morphology – White Matter Composition Correlation in Patients with NF1

2D morphological measurements do not distinguish between grey or white matter composition of the structures measured. In order to determine whether the increased CC and brainstem measurements we observed were related to altered myelination (white matter), we correlated 2D brain morphological measurements with 3D brain volumetric measurements in individuals with NF1. Principal component analysis was used to identify combinations of CC or brainstem 2D measurements that distinguished NF1 patients. Ninety-nine adults with NF1 (47 females and 52 males) had complete 2D and 3D measurements available for the CC. The first principal component (PC1) reflects a combination of CC area and mid-body measurements so that a larger PC1 indicates greater CC area and mid-body width. Larger PC2 indicates shorter and flatter CC. Larger PC3 indicates less bulbous CC posterior body. Larger PC4 indicates male. Overall, we found that larger 2D corpus callosum measurements most strongly correlated with increased total brain volume and increased corpus callosum white matter volume (Figure 4A).

**Figure 4:**
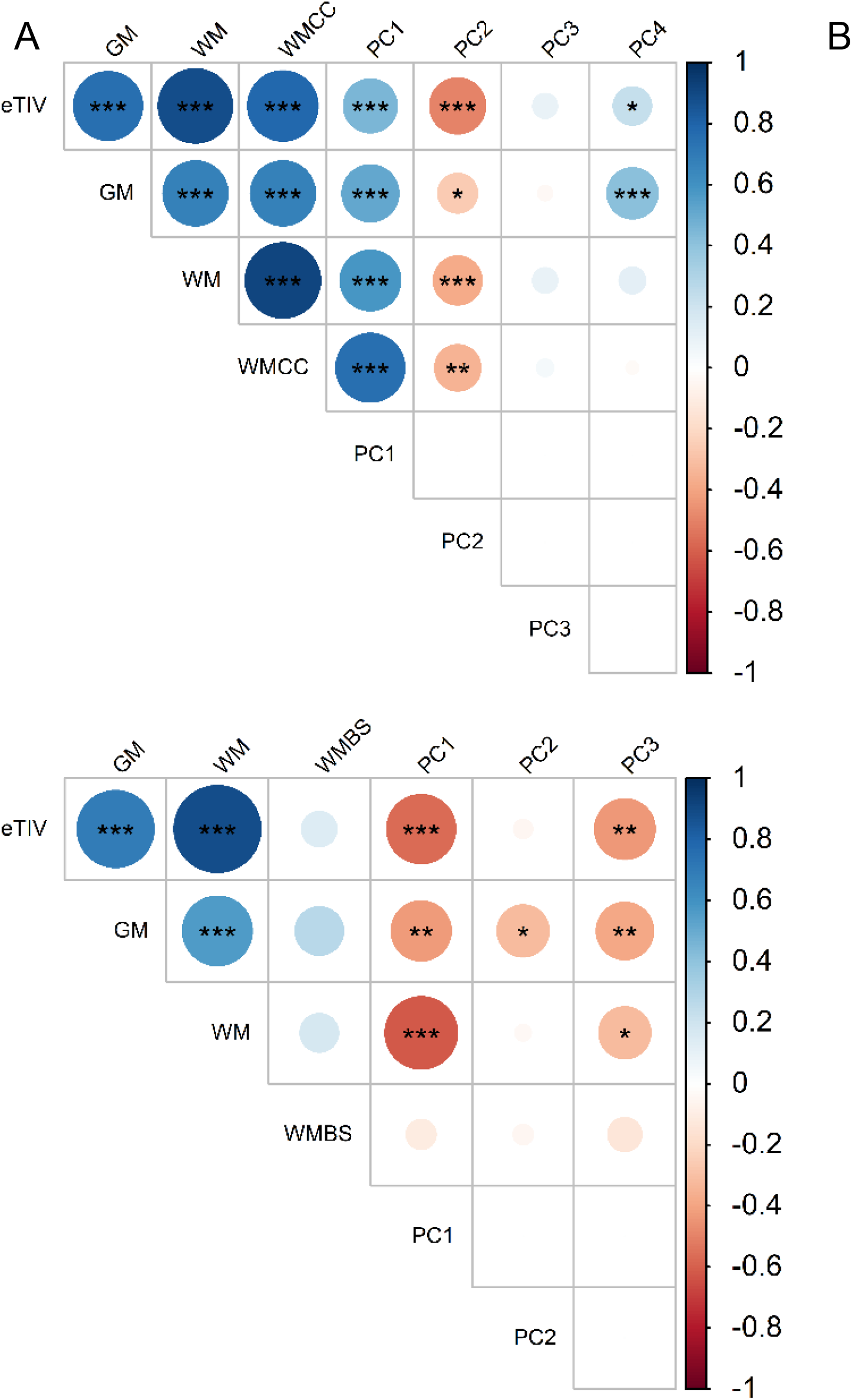
Correlation matrix of principal components of 2D brain morphological measurements with 3D brain volumetric measurements of the corpus callosum (A) and brainstem (B) in adults with NF1. The color scale represents the degree of correlation from strongly negative (−1, red) to strongly positive (1, blue). The size of the circles also indicates the strength of the correlation, with values closer to -1 or 1 larger than those that are closer to 0. Asterisks indicate FDR-adjusted statistical significance. p < 0.05 = *, p< 0.01 = **, p < 0.001 = ***. Abbreviations: eTIV = total brain volume, GM = grey matter volume, WM = white matter volume, WMCC = corpus callosum white matter volume, WMBS = brainstem white matter volume, and PC = principal component.

There were 55 individuals with NF1 (25 females and 30 males) who had complete 2D and 3D measurements for the brainstem. The top three PCs based on 2D brainstem measurements were selected for correlation analysis. Larger PC1 indicates smaller pons and MCP, larger PC2 indicates older age, and larger PC3 indicates female. We found that larger 2D pons and MCO measurements most strongly correlate with increased total brain volume and total white matter volume (Figure 4B).

These results suggest that the greater than expected 2D corpus callosum and brainstem measurements reflect increased total white matter volume, and, by extension, myelination.

### Morphology - Neuropsychometric Correlations

Twenty-nine adults with NF1 had complete datasets for the clinical and neuropsychometric assessments as well as for the CC measurements analysed. The top three PCs were selected for the CC. None of the CC PCs was strongly correlated with any of the five clinical assessments or neuropsychometric measurements (Table 2).

**Table 2:** Correlation values and p-values of corpus callosum measurements with clinical and neuropsychometric assessments in adults with NF1.

| CORRELATION MATRIX |  |  |  |  |  |  |  |  |
| --- | --- | --- | --- | --- | --- | --- | --- | --- |
|  | Clin. Sev. | Neuro. Sev. | ADD/ADHD | IQ | ACS | PC1 | PC2 | PC3 |
| Clin. Sev. |  | *0.14* | *-0.49* | *-0.29* | *-0.17* | *0.38* | *-0.05* | *-0.02* |
| Neuro. Sev. |  |  | *0.55* | *0.20* | *-0.41* | *0.19* | *-0.05* | *0.17* |
| ADD/ADHD |  |  |  | *0.26* | *-0.17* | *0.01* | *-0.20* | *-0.06* |
| IQ |  |  |  |  | *0.10* | *-0.01* | *-0.41* | *0.33* |
| ACS |  |  |  |  |  | *-0.22* | *0.23* | *0.08* |
| PC1 |  |  |  |  |  |  | *-0.20* | *-0.03* |
| PC2 |  |  |  |  |  |  |  | *-0.09* |

Top three corpus callosum PCs are correlated to neuropsychometric measurements (clinical severity, clinical neurological severity, clinical ADD/ADHD diagnosis, IQ, and Attention Comparison Score). The correlation matrix combines the Pearson correlation and Spearman correlation (italicized) results. Abbreviations used in the table: Clin. Sev. = clinical NF1 severity, Neuro. Sev. = clinical neurological severity, ADD/ADHD = clinical Attention deficit hyperactivity disorder severity, IQ = intelligence quotient, ACS = Attention Comparison Score, and PC = principal component.

Twenty-six adults with NF1 had complete datasets for the clinical and neuropsychometric assessments and brainstem measurements analysed. The top three PCs were selected for the brainstem. PC1 was correlated with both decreased neurological severity (*r*_*s*_ = -0.51) and negative clinical ADD/ADHD diagnosis (*r*_*s*_ = -0.49). The three largest weights for PC1 were the right MCP length (−0.42), left MCP length (−0.41), and the pons AP length (−0.40). Thus, a smaller PC1 value indicates an overall thicker middle brainstem. Clinical severity was correlated with decreased clinical ADD/ADHD diagnosis (*r*_*s*_ = -0.56), and neurological severity was correlated with decreased IQ (*r*_*s*_ = -0.38) (Table 3).

**Table 3:** Correlation values and p-values of brainstem measurements with clinical and neuropsychometric assessments in adults with NF1.

| CORRELATION MATRIX |  |  |  |  |  |  |  |  |
| --- | --- | --- | --- | --- | --- | --- | --- | --- |
|  | Clin. Sev. | Neuro. Sev. | ADD/ADHD | IQ | ACS | PC1 | PC2 | PC3 |
| Clin. Sev. |  | *-0.06* | *-0.56* | *-0.24* | *-0.13* | *0.21* | *-0.17* | *0.21* |
| Neuro. Sev. |  |  | *0.43* | *-0.38* | *-0.21* | *-0.51* | *-0.16* | *-0.20* |
| ADD/ADHD |  |  |  | *0.24* | *-0.06* | *-0.49* | *0.32* | *-0.31* |
| IQ |  |  |  |  | *0.35* | *-0.17* | *0.33* | *0.17* |
| ACS |  |  |  |  |  | *0.05* | *0.27* | *0.30* |
| PC1 |  |  |  |  |  |  | *0.01* | *0.18* |
| PC2 |  |  |  |  |  |  |  | *0.22* |

Top three brainstem PCs are correlated to neuropsychometric measurements (clinical severity, clinical neurological severity, clinical ADD/ADHD diagnosis, IQ, and Attention Comparison Score). The correlation matrix combines the Pearson correlation and Spearman correlation (italicized) results. Abbreviations used in the table: Clin. Sev. = clinical NF1 severity, Neuro. Sev. = clinical neurological severity, ADD/ADHD = clinical Attention deficit hyperactivity disorder severity, IQ = intelligence quotient, ACS = Attention Comparison Score, and PC = principal component.

Forty-four adults with NF1 had complete datasets for the clinical and neuropsychometric assessments and ocular globe measurements analysed. The top five PCs were selected for the ocular globes. None of the variables was strongly correlated with the others (Table 4).

**Table 4:** Correlation values and p-values of ocular globe measurements with clinical and neuropsychometric assessments in adults with NF1.

| CORRELATION MATRIX |  |  |  |  |  |  |  |  |  |  |
| --- | --- | --- | --- | --- | --- | --- | --- | --- | --- | --- |
|  | Clin. Sev. | Neuro. Sev. | ADD/ADHD | IQ | ACS | PC1 | PC2 | PC3 | PC4 | PC5 |
| Clin. Sev. |  | *0.23* | *-0.21* | *-0.21* | *-0.21* | *-0.36* | *0.01* | *0.04* | *-0.40* | *-0.20* |
| Neuro. Sev. |  |  | *0.49* | *-0.34* | *-0.13* | *-0.25* | *0.20* | *-0.08* | *0.23* | *0.11* |
| ADD/ADHD |  |  |  | *0.04* | *-0.03* | *0.13* | *0.36* | *-0.18* | *0.10* | *0.26* |
| IQ |  |  |  |  | *0.16* | *0.50* | *-0.11* | *-0.01* | *-0.02* | *-0.36* |
| ACS |  |  |  |  |  | *-0.10* | *-0.18* | *-0.07* | *0.09* | *0.01* |
| PC1 |  |  |  |  |  |  | *0.12* | *0.06* | *0.08* | *-0.05* |
| PC2 |  |  |  |  |  |  |  | *0.06* | *-0.04* | *0.04* |
| PC3 |  |  |  |  |  |  |  |  | *-0.05* | *0.04* |
| PC4 |  |  |  |  |  |  |  |  |  | *0.00* |

Top five corpus callosum PCs are correlated to neuropsychometric measurements (clinical severity, clinical neurological severity, clinical ADD/ADHD diagnosis, IQ, and Attention Comparison Score). The correlation matrix combines the Pearson correlation and Spearman correlation (italicized) results. Abbreviations used in the table: Clin. Sev. = clinical NF1 severity, Neuro. Sev. = clinical neurological severity, ADD/ADHD = clinical Attention deficit hyperactivity disorder severity, IQ = intelligence quotient, ACS = Attention Comparison Score, and PC = principal component.

## DISCUSSION

We conducted the first large-scale MRI study of brain morphological differences and their relationship to cognitive or behavioural abnormalities in adults with NF1. We found that adults with NF1 have apparent enlargement of the corpus callosum and brainstem in comparison to unaffected adults. These 2D morphological enlargements are correlated to increased total white matter volume. In our companion study focused on brain volume, we also found an increase in total and regional white matter in the brains of adults with NF1 compared to control adults^28^. In the current study, we did not find any obvious correlation between the brain morphological changes we observed and the clinical or neuropsychometric assessments in these individuals.

In the present study, we observed increased CC area, height, and anterior body and mid-body widths among adults with NF1. Our findings are consistent with those of previous studies in children and smaller groups of adults with NF1^23, 25^. Also, previous brain MRI studies in people with NF1 have found evidence of increased total white matter volume, increased brain volume, and megalencephaly^6, 7, 9, 10, 19-22^. Similarly, we found strong correlation between 2D CC size and total and CC white matter volume.

Loss of function mutations of *NF1* cause dysregulation of proliferation in Schwann cells, which are responsible for the myelination of axons in the peripheral nervous system^16^. In the brain, white matter is mainly comprised of myelinated axons^39^. It is hypothesized that the increases in total white matter volume, total brain volume, and megalencephaly that occur in NF1 are related to the dysregulation of oligodendrocytes, the myelin-producing cells in the central nervous system^23, 40^. If this interpretation is correct, enlargement of the CC, a structure largely composed of white matter, might be expected in people with NF1. The decrease in genu width and increase in the anterior and mid-body widths raises the possibility that the CC shape is also changed. Enlargement of the CC height and area without significant alteration of the length is consistent with a change in CC shape and volume.

The enlargement of the brainstem that we observed among adults with NF1 is a novel finding. The brainstem consists of both grey and white matter, with the MCP (which was clearly enlarged among adults with NF1) comprised mostly of white matter^41, 42^. This is further evident by the strong correlation we found between increased brainstem 2D morphology measurements (specifically pons and MCP) and increased total white matter volume. Thus, the brainstem enlargement we observed is also consistent with dysregulated myelin proliferation in individuals with NF1.

Both age and sex are known to affect CC size, ocular globe size and position, and brainstem size^43-47^. We carefully matched unaffected individuals by age and sex to avoid confounding by these factors in our analysis. However, the adult NF1 group was recruited from Hamburg, Germany, while the unaffected comparison group was obtained in Vancouver, Canada. Different MR imaging procedures and measurement software were used for the NF1 group and the unaffected comparison control group. The different imaging procedures resulted in an inability to detect differences between the two groups smaller than 0.05 cm, but all statistically significant differences in brain morphological measurements observed between NF1 and control subjects were greater than 0.05 cm.

Another limitation of our study is the lack of dedicated orbital MRIs. The asymmetry in the ONT we observed probably does not represent a true anatomical difference and may be a result of measurement error, as previous studies have not found ONT asymmetry^26, 27^. The current study is less accurate than Ji et al.’s (2013), as we only measured ONT in one axial plane while Ji and associates used dedicated 3-dimensional magnetization-prepared rapid gradient echo sequences with 1 mm slices^27^.

Our correlation analyses between structures measured on MRI and clinical/neuropsychometric assessments are limited by small sample sizes as only a subset of the adults with NF1 had psychometric testing. Some previous studies have found that increased CC volume or CC index correlated with decreased academic achievement and IQ in children with NF1^10, 24^. In contrast, Kayl et al. (2000b) found that among children with NF1, smaller splenium size was correlated with increased attention problems as reported by teachers^29^. Furthermore, a more recent study using diffusion tensor imaging to examine myelination of white matter specifically failed to find a significant relationship between total CC area and IQ scores in children with NF1^30^. We did not find any robust correlations between the brain morphology and neuropsychometric measurements in adults with NF1.

Clinically, 2D MRI is used more often than 3D MRI. We conducted the largest study of 2D brain morphology in adults with NF1 reported to date to characterize the brain morphology alterations. The enlargement of the CC and brainstem and correlation to increased total white matter volume that we observed in adults with NF1 lends support to the hypothesis that neurofibromin haploinsufficiency causes dysregulation of myelin production in the brain. The relationship of this overgrowth of myelinated brain structures to the frequent occurrence of central nervous system gliomas and of benign and malignant peripheral nerve sheath tumours in individuals with NF1 is unknown but merits further study.

## Data Availability

Anonymous data is available for appropriate research purposes through V.F. Mautner, MD.

## ACKNOWLEDGEMENT

We would like to thank Sofia Granstroem who analysed the neuropsychometric data. We are grateful to the Bundesverband Neurofibromatose organization for funding this project.

## AUTHOR CONTRIBUTIONS

SW contributed to design of the study, collected all 2D MRI morphological data, analyzed all data, and drafted and revised manuscript. VFM contributed to conception and design of the study, recruited patients, played a major role in the acquisition of MRI and neuropsychometric data, and revised the manuscript for intellectual content. RB contributed to the acquisition and analysis of volumetric MRI data, and revised manuscript for intellectual content. SF advised on key statistical analyses and revised the manuscript for intellectual content. PS conducted the volumetric MRI analyses and revised the manuscript for intellectual content. JMF contributed to conception and design of the study and revised the manuscript for intellectual content. MKSH contributed to design of the study, guided morphological data collection, and revised the manuscript for intellectual content. JMF and MKSH contributed equally to the manuscript.

